# Applications of Large Language Models (LLMs) in Breast Cancer Care

**DOI:** 10.1101/2023.11.04.23298081

**Authors:** Vera Sorin, Benjamin S. Glicksberg, Yiftach Barash, Eli Konen, Girish Nadkarni, Eyal Klang

## Abstract

**Purpose:** Recently introduced Large Language Models (LLMs) such as ChatGPT have already shown promising results in natural language processing in healthcare. The aim of this study is to systematically review the literature on the applications of LLMs in breast cancer diagnosis and care.

**Methods:** A literature search was conducted using MEDLINE, focusing on studies published up to October 22nd, 2023, using the following terms: “large language models”, “LLM”, “GPT”, “ChatGPT”, “OpenAI”, and “breast”.

**Results:** Five studies met our inclusion criteria. All studies were published in 2023, focusing on ChatGPT-3.5 or GPT-4 by OpenAI. Applications included information extraction from clinical notes, question-answering based on guidelines, and patients’ management recommendations. The rate of correct answers varied from 64-98%, with the highest accuracy (88-98%) observed in information extraction and question-answering tasks. Notably, most studies utilized real patient data rather than data sourced from the internet. Limitations included inconsistent accuracy, prompt sensitivity, and overlooked clinical details, highlighting areas for cautious LLM integration into clinical practice.

**Conclusion:** LLMs demonstrate promise in text analysis tasks related to breast cancer care, including information extraction and guideline-based question-answering. However, variations in accuracy and the occurrence of erroneous outputs necessitate validation and oversight. Future works should focus on improving reliability of LLMs within clinical workflow.

## Introduction

Natural language processing (NLP) is increasingly being used in healthcare, particularly within oncology, allowing free-text analysis, with various applications^1^. This advancement has been further amplified by the recent advent of large language models (LLMs). LLMs such as GPT, LLaMA, PaLM, and Falcon, are deep learning NLP algorithms^2^ that are based on the transformer architecture. They are composed of billions of parameters, enabling processing and generation of text with remarkable accuracy^3^. Research into healthcare applications of these models is expanding^4-8^. GPT-4, for instance, has achieved an 87% success rate on the USMLE^9,10^. With recent developments, it can now also be applied to image analysis^11^.

Breast cancer stands as the most common cancer among women, leading to significant morbidity, mortality, and widespread concern^6,12^. With the increasing volume of medical data available, both clinicians and patients face the challenge of navigating and interpreting vast amounts of information. In this context, LLM technology can be helpful, enabling automatic processing and presenting of relevant data. Recent studies have evaluated applications of LLMs in breast cancer diagnosis and management.

The aim of this study is to review the literature on applications of LLMs in breast cancer care.

## Methods

We conducted a comprehensive literature search on the applications of LLMs in breast cancer diagnosis and care using MEDLINE. The search included studies published up to October 22^nd^ 2023. Our search query was “((“large language models”) OR (llm) OR (gpt) OR (chatgpt) OR (openAI)) AND (breast)”. The initial search identified 96 studies. To ensure thoroughness, we also examined the reference lists of the relevant studies. This however did not lead to additional relevant studies that met our inclusion criteria.

The criteria for inclusion in our review were English language full-length publications that specifically evaluated the role and impact of LLMs in breast cancer diagnosis and care. We excluded papers that addressed other general applications of LLMs in healthcare or oncology without a specific focus on breast cancer diagnosis and care.

Two reviewers (VS, EKL) independently conducted the search, screened the titles, and reviewed the abstracts of the articles identified in the search. One discrepancy in the search results was discussed and resolved to achieve a consensus. Following this, the reviewers assessed the full text of the relevant papers. In total, five publications met our criteria and were incorporated into this review. We summarized the results of the included studies, detailing the specific LLMs used, the utilized tasks, number of cases, along with publication details in a table format. **Figure 1** provides a flowchart detailing the screening and inclusion procedure.

**Figure 1.**
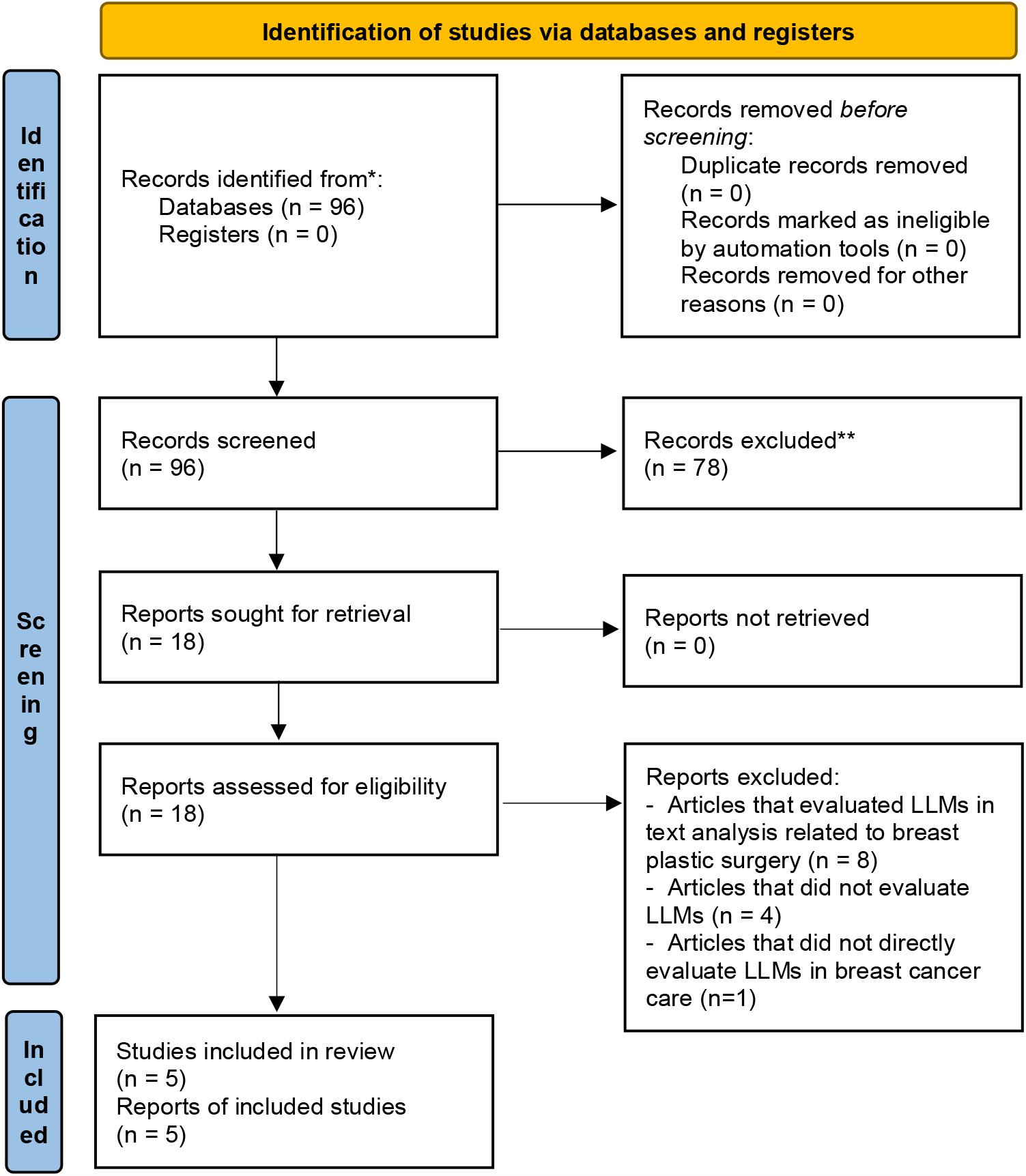
Flow Diagram of the Inclusion Process Flow diagram of the search and inclusion process based on the Preferred Reporting Items for Systematic Reviews and Meta-Analyses (PRISMA) guidelines

## Results

All five studies included in this review were published in 2023 (**Table 1**). All studies focused on either ChatGPT-3.5 or GPT-4 by OpenAI. Applications described include information extraction and question-answering. Three studies (60.0%) evaluated the performance of ChatGPT on actual patient data^13-15^, as opposed to two studies that used data from the internet^16,17^.

**Table 1.** Studies Evaluating LLMs for Breast Cancer Diagnosis and Care.

| **Study** <sup>ref.</sup> | **Publication Date** | **Title** | **Journal** |
| --- | --- | --- | --- |
| Sorin et al. <sup>13</sup> | 05.2023 | Large language model (ChatGPT) as a support tool for breast tumor board | NPJ Breast Cancer |
| Rao et al. <sup>16</sup> | 06.2023 | Evaluating GPT as an Adjunct for Radiologic Decision Making: GPT-4 Versus GPT-3.5 in a Breast Imaging Pilot | JACR |
| Choi et al. <sup>14</sup> | 09.2023 | Developing prompts from large language model for extracting clinical information from pathology and ultrasound reports in breast cancer | Radiation Oncology Journal |
| Lukac et al. <sup>15</sup> | 07.2023 | Evaluating ChatGPT as an adjunct for the multidisciplinary tumor board decision-making in primary breast cancer cases | Archives of Gynecology and Obstetrics |
| Haver et al. <sup>13</sup> | 04.2023 | Appropriateness of Breast Cancer Prevention and Screening Recommendations Provided by ChatGPT | Radiology |

Rao et al. and Haver et al. evaluated LLMs for breast imaging recommendations^16,17^, Sorin et al. and Lukac et al. evaluated LLMs as supportive decision making tools in multidisciplinary tumor boards^13,15^, and Choi et al. used LLM for information extraction from ultrasound and pathology reports^14^, (**Figure 2**). Performance of LLMs on different applications ranged from 64-98%. Best performance rates were achieved for information extraction and question-answering, with correct responses ranging from 88-98%^14,16^ (**Table 2**).

**Table 2.** Summarization of Performance of LLMs at Different Breast Cancer Care Related Tasks.

| **Study**<sup>ref.</sup> | **LLM** | **No. of Cases** | **Actual Patient Data** | **Application** | **Correct Performance** |
| --- | --- | --- | --- | --- | --- |
| Sorin et al.<sup>13</sup> | ChatGPT (GPT-3.5) | 10 | Yes | Tumor board clinical decision support | 70% |
| Rao et al.<sup>16</sup> | GPT-4, GPT-3.5 | 14 | No | Question-answering based on ACR recommendations | 88.9% - 98.4% |
| Choi et al.<sup>14</sup> | ChatGPT (GPT-3.5) | 340 | Yes | Information extraction | 87.7% - 98.2% |
| Lukac et al.<sup>15</sup> | ChatGPT (GPT-3.5) | 10 | Yes | Tumor board clinical decision support | 64.20% |
| Haver et al.<sup>17</sup> | ChatGPT (GPT-3.5) | 25 | No | Question-answering on breast cancer prevention and screening | 88% |

**Figure 2.**
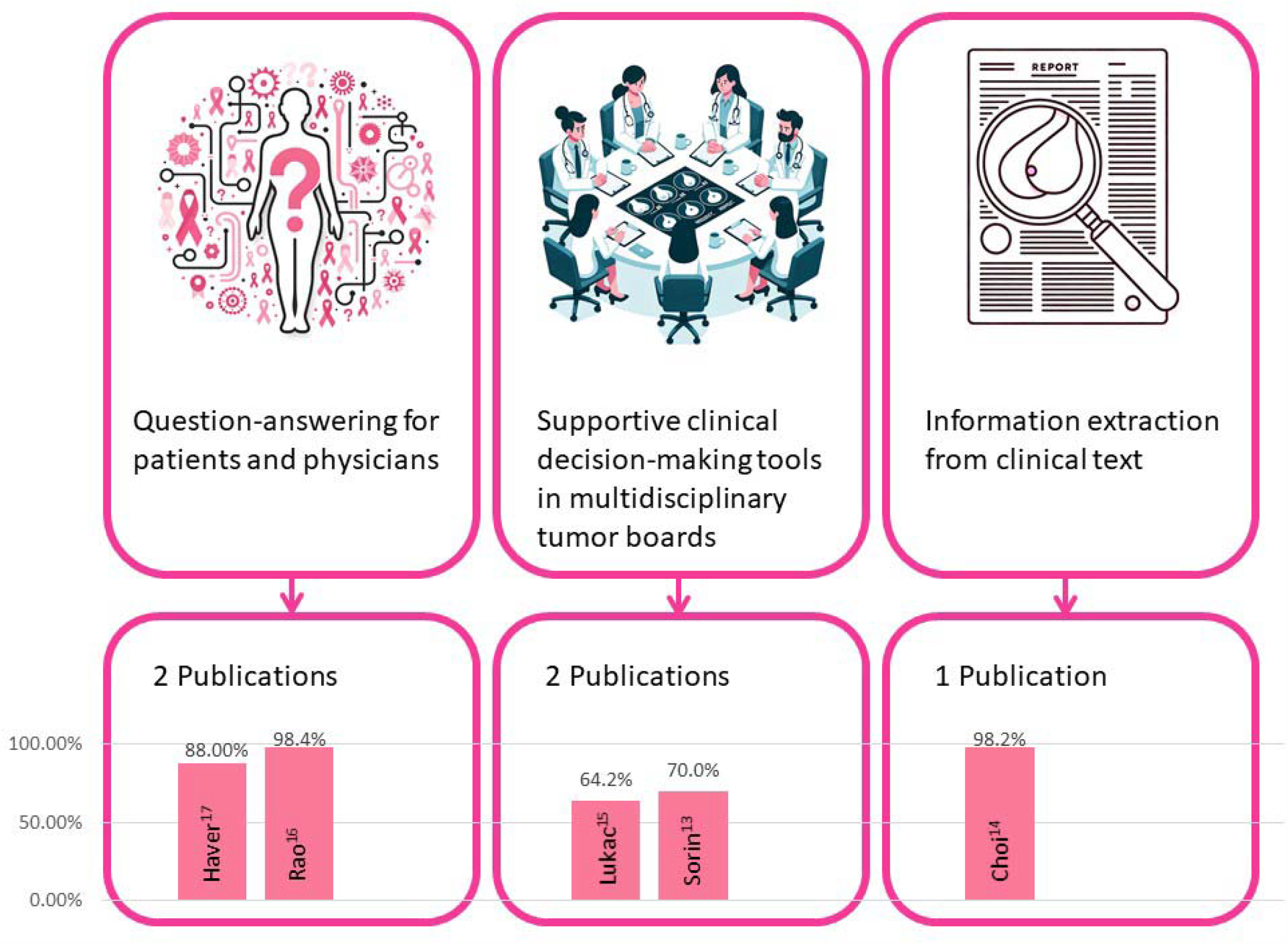
Applications of large language models in breast cancer care and the corresponding accuracies achieved in various tasks in the different studies

All studies discussed limitations of LLMs in the contexts the algorithms were evaluated **(Table 3)**. In all studies some of the answers and information the models generated was false. When used as a support tool for tumor board, in some instances, the models overlooked relevant clinical details^13,15^. Sorin et al. noticed absolute lack of referral to imaging^13^, while Rao et al. who evaluated appropriateness of imaging noticed imaging overutilization^16^. Some of the studies also discussed prompt sensitivity^14,17^, and difficulty to verify the reliability of the answers^15-17^.

**Table 3.** Limitations of LLMs as Described in Each Study.

| **Study**<sup>ref.</sup> | **LLM** | **Limitations Described** |
| --- | --- | --- |
| Sorin et al.<sup>13</sup> | ChatGPT (GPT-3.5) | False answers and inaccurate medical recommendations, overlooked relevant clinical details, absolute lack of referral to imaging, potential for outdated information, potential for bias |
| Rao et al.<sup>16</sup> | GPT-4, GPT-3.5 | False information, imaging overutilization, lack of source attribution |
| Choi et al.<sup>14</sup> | ChatGPT (GPT-3.5) | False information, lack of logical reasoning, incomplete information extraction, prompt sensitivity |
| Lukac et al.<sup>15</sup> | ChatGPT (GPT-3.5) | False answers, overlooked relevant clinical details, potential for outdated information, lack of source attribution |
| Haver et al.<sup>17</sup> | ChatGPT (GPT-3.5) | False recommendations, prompt sensitivity, lack of source attribution |

## Discussion

In this study we reviewed the literature on LLMs applications for breast cancer diagnosis and care. Applications described included information extraction from clinical texts, question-answering for patients and physicians, manuscript drafting and clinical management recommendations.

Performance ranged from 64-98% correct answers generated by the LLM, with best performance in question answering and information extraction tasks.

Interestingly, most studies in this review included real patients’ data as opposed to data from the internet. When looking at the overall published literature on LLMs applications in healthcare, there are more publications evaluating LLMs performance on data from the internet, including performance on board examinations and question-answering based on guidelines^4^. These analyses may introduce contamination of data during model training, owing to the fact that LLMs were trained on vast data from the internet. For commercial models such as OpenAI’s ChatGPT, the type of training data is not disclosed. Furthermore, these applications do not necessarily reflect on the performance of these models in real-world clinical setting.

The variety of tasks described in this review highlight the potential of LLMs in text analysis related to breast cancer care. However, while some claim that these models may eventually replace healthcare personnel, currently, there are major limitations and ethical concerns that will not allow this^18^. Using such models to augment physicians’ performance is more practical, albeit also constrained by ethical issues^19^. LLMs enable automating different tasks that traditionally required human effort. An ability to analyze, extract and generate meaningful textual information could potentially decrease some of physicians’ workload and perhaps even decrease human errors.

The reliance on LLMs and their potential integration in medicine should be balanced with caution. The limitations discussed in the studies further underscore this note. These models can generate false information (termed “hallucination”) which can be seamlessly and confidently integrated into real information^1^. They can also perpetuate disparities in healthcare^20,21^. The inherent inability to trace the exact decision-making process of these algorithms is a major challenge for trust and clinical integration^22^. These models can also be vulnerable to cyber-attacks^23^.

This review has several limitations. First, due to the heterogeneity of tasks evaluated in the studies, we could not perform a meta-analysis. Second, we only included studies evaluating breast cancer related data. There are many studies that evaluate applications in oncology that may be relevant and extend to examples including breast cancer patients, these were not included. Third, all included studies assessed ChatGPT-3.5, and only one study evaluated GPT-4. There were no publications identified on other available LLMs. Finally, generative AI is currently a rapidly expanding topic. Thus, there may be manuscripts and applications published after our review was performed. LLMs are continually being refined, and so is their performance.

To conclude, LLMs show promise in text analysis related to breast cancer care, enabling information extraction and guideline-based question-answering. However, variations in accuracy and the occurrence of erroneous outputs necessitate validation and oversight. Future work should focus on improving the reliability of LLMs within clinical workflow.

## Data Availability

All data produced in the present work are contained in the manuscript

